# Implementation of a specialist pneumonia intervention nurse service significantly lowers mortality for community acquired pneumonia

**DOI:** 10.1101/19007336

**Authors:** Robert C. Free, Matthew Richardson, Camilla Pillay, Julie Skeemer, Kayleigh Hawkes, Rebecca Broughton, Pranabashis Haldar, Gerrit Woltmann

## Abstract

**Objectives:** Evaluate clinical outcomes associated with implementing a specialist pneumonia intervention nursing (SPIN) service, to improve adherence with BTS guidelines for hospitalised community acquired pneumonia (CAP).

**Design:** Retrospective cohort study, comparing periods before (2011-13) and after (2014-16) SPIN service implementation.

**Setting:** Single NHS trust across two hospital sites in Leicester City, England

**Participants:** 13,496 adult (aged ≥16) admissions to hospital with a primary diagnosis of CAP

**Interventions:** The SPIN service was set up in 2013/2014 to provide clinical review of new CAP admissions; assurance of guidelines adherence; delivery of CAP clinical education and clinical follow up after discharge.

**Main outcome measures:** The primary outcomes were proportions of CAP cases receiving antibiotic treatment within 4 hours of admission and change in crude in-hospital mortality rate. Secondary outcomes were adjusted mortality rate and length of stay (LOS).

**Results:** The SPIN service reviewed 38% of CAP admissions in 2014-16. 82% of these admissions received antibiotic treatment in <4 hours (68.5% in the national audit). Compared with the pre-SPIN period, there was a significant reduction in both 30-day (OR=0.77 [0.70-0.85], p<0.0001) and in-hospital mortality (OR=0.66 [0.60-0.73], p<0.0001) after service implementation, with a review by the service having the largest independent 30-day mortality benefit (HR=0.60 [0.53-0.67], p<0.0001). There was no change in LOS (median 6 days).

**Conclusions:** Implementation of a SPIN service improves adherence with BTS guidelines and achieves significant reductions in CAP associated mortality. This enhanced model of care is low cost, highly effective and readily adoptable in secondary care.

**Key Messages:** *What is the key question?:* Does a specialist nurse-led intervention affect BTS guideline adherence and mortality for patients admitted to hospital with community acquired pneumonia (CAP)?

*What is the bottom line?:* Implementing specialist nurse teams for CAP delivers improved guideline adherence and survival for patients admitted with the condition.

*Why read on?:* This study shows a low-cost specialist nursing service focussed on CAP is associated with a significant improvement in BTS guidelines adherence and patient survival.

## Introduction

Community acquired pneumonia (CAP) is the leading cause of in-hospital mortality, with a crude rate of 5-15%, rising to 30% for in patients who are admitted to ICU [1]. Pneumonia is responsible for more hospital admissions and bed days than any other lung disease in the UK and annual healthcare costs to the NHS associated with CAP are estimated to exceed £1 billion. Despite this, pneumonia has historically been a substantially underestimated, frequently neglected and underfunded condition in the UK. This was recognised in the recently published NHS Long Term Plan [2] which highlights CAP as an NHS research priority for new treatments and care pathways.

CAP mortality is known to be associated with disease severity and a number of scores have been developed to quantify this, of which CURB-65 [3] is implemented most widely in practice. These scores are intended to improve the provision of prompt and appropriate clinical care for pneumonia and their benefit for guiding clinical intervention has been demonstrated in a national implementation project that reported lower 30 day in-hospital mortality associated with prompt radiological diagnosis and severity based treatment [4,5]. This suggests that prognosis in CAP is modifiable and can be improved with better models of healthcare delivery.

University Hospitals Leicester NHS Trust (UHL) provides emergency admissions facilities for a catchment population of 1 million people. The Trust previously registered higher than expected externally reported Summary Hospital-level Mortality Indicators (SHMI) [6] associated with primary diagnosis of CAP. In response, we created a dedicated specialist pneumonia intervention nursing (SPIN) service in 2013/14 with the aims of i) improving systematic CURB-65 scoring for CAP admissions; and ii) improving and accelerating adherence to key components of the BTS pneumonia guidelines. This included radiological diagnosis and prompt antibiotic treatment within 4 hours of admission. In addition, the SPIN team led a trust-wide educational and awareness-raising campaign on the importance of guidelines based management of acute CAP cases and facilitated radiological follow up after discharge.

In this five year retrospective cohort study, comparing periods before, during and after implementation of the SPIN service, we report on the effectiveness of this nurse led programme of interventions for improving guidelines adherence and the impact of the SPIN service on crude in-hospital mortality, SHMI and HSMR (Hospital Standardized Mortality Ratio).

## Methods

This manuscript follows the Standards for Quality Improvement Reporting Excellence 2.0 guidelines for study design and analysis [7].

### The SPIN Team

In 2013/14 a new SPIN service was created, to which two nurses were appointed and tasked with implementation of key measures to improve diagnosis and prompt management of CAP admissions. The nurse posts were funded through the Commissioning for Quality and Innovation payments framework for two years before long term funding was secured.

An assessment checklist based on the BTS pneumonia guidelines was devised (Supplementary File 1). This checklist was printed on adhesive stickers for the medical notes and served as an educational resource for awareness raising during the implementation period. Embedded management guidance was disseminated widely using wall posters in admission areas, the hospital intranet and as part of dedicated education modules delivered by the team. The service was operational weekdays between 9am and 5pm.

Eligible emergency admissions were identified from daily ward list screening, review of admission chest x-rays on the picture archiving and communication system and in response to electronic or verbal referral of patients with CAP (Figure 1). Cases were predominantly seen in acute medical admission areas at both UHL emergency sites (Leicester Royal Infirmary and Glenfield Hospital). Severity scores (CURB-65) and interventions were prospectively recorded and time stamped for admissions physically seen by the SPIN team using bespoke intranet database facilities (NIHR Biomedical Research Centre - Respiratory, Leicester).

**Figure 1:**
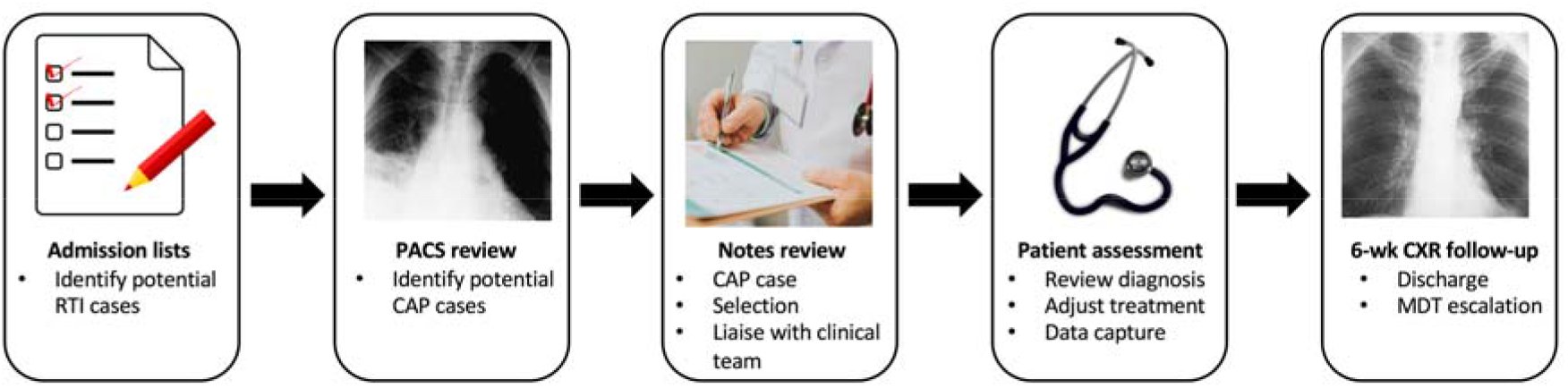
Schematic for admission screening of CAP patients.

### Data extraction and analysis

Data across five financial years (FY) were categorised into three periods for comparative analysis: i) a two year baseline period (FY 2011/12 - 2012/13) prior to availability of the SPIN service; ii) a one year implementation phase (FY 2013/14) during which the SPIN service was set up and operationalised; and iii) a two year intervention period (FY 2014/15 - 2015/16) when SPIN services were fully operational. This period was stratified further according to cases seen or not seen by the nursing service.

To ensure UHL admissions data was representative and unaffected by clinical coding bias, comparative per-hospital trust admission frequency data was obtained from Telstra/Dr Foster.

Individual CAP coded hospital episodes from the study period were extracted from the hospital data warehouse by adapting the algorithm used by Telstra/Dr Foster for their quality and outcomes analysis. The International Classification of Diseases (ICD-10) clinical codes representing pneumonia in the Clinical Classifications Software [8] group were used (Supplementary File 2) for this purpose. A patient and date/time matching algorithm linked CAP database intervention data to coded admission records, prior to anonymisation for analysis. Additional data fields derived from the anonymised admissions data were length of stay (LOS), Charlson comorbidity index (CCI) [6,9], and time to death in days and weeks. CCI and age were categorised into groups of 0, 1-5 and >5, and 16 - 65 and >65, respectively. The presence of heart disease and/or diabetes comorbidities in admissions were determined by comparing secondary diagnoses with ICD-10 code lists (Supplementary File 3).

All statistical analyses were performed using R version 3.3.1 [10]. Comparisons between baseline and intervention variables were performed using the chi-square test, with Wald’s method used to calculate 95% confidence intervals. SHMI and HSMR adjusted outcome data were provided by Telstra/Dr Foster and included alongside the results.

Survival analysis utilised Kaplan-Meir (KM) curves and Cox proportional hazards (PH) models. Patients were censored if they had died after the one-year period examined or if they had survived beyond this time. KM survival analysis was carried out between baseline and intervention groups using time to death. A Cox PH model was constructed including potential confounding variables. Male gender and weekend admission were included as binary variables, age and CCI as continuous variables.

#### Patient and Public Involvement

The investigation did not include PPI in the design, analysis, or interpretation of this data. The analysis was done using anonymised data, and therefore we were unable to work with participants or disseminate the results to them.

## Results

### Descriptive CAP admission statistics

There were 13,496 CAP admissions to UHL between FY 2011 - 2016. The median age of admissions was 77.0 (IQR 64.0 - 86.0) years. Compared with the baseline period, there was an increase during the intervention period in the number of hospital admissions for CAP (4,143 vs 7,029). Standardised stationary series comparing monthly local CAP admission frequency at UHL with UK frequencies demonstrated concordance in the trend and seasonality of cases (Supplementary File 4). The rise in CAP admissions during the intervention period was accompanied by an increase in the proportion of patients with CAP aged >65 years (72.0% vs 73.8%, OR=1.09 [1.00-1.19], p<0.046) and the proportion with complex co-morbidities (CCI >5) (from 41.9% to 47.8%, OR=1.27 [1.18-1.38], p<0.0001) (Table 1).

**Table 1:** Comparison of admissions between baseline and SPIN intervention periods

|  | Baseline<br>% (n = 4143) | SPIN intervention<br>% (n = 7029) | OR | 95% CI | p |
| --- | --- | --- | --- | --- | --- |
| **Male** | 52.1 (2159) | 50.7 (3565) | 0.95 | 0.88 - 1.02 | 0.155 |
| **Age** |  |  |  |  |  |
| 16-65 | 28.0 (1158) | 26.2 (1846) | 0.92 | 0.84 - 1.00 | **0.046** |
| >65 | 72.0 (2985) | 73.8 (5187) | 1.09 | 1.00 - 1.19 | **0.046** |
| **Charlson**<br>**Comorbidity**<br>**Index** |  |  |  |  |  |
| 0 | 34.8 (1443) | 28.8 (2024) | 0.76 | 0.70 - 0.82 | **<0.0001** |
| 1-5 | 23.3 (966) | 23.4 (1642) | 1.00 | 0.92 - 1.10 | 0.957 |
| >5 | 41.9 (1734) | 47.8 (3363) | 1.27 | 1.18 - 1.38 | **<0.0001** |
| **Weekend**<br>**(Sat/Sun)** | 26.6 (1103) | 27.8 (1953) | 1.06 | 0.97 - 1.16 | 0.183 |

### Successful implementation and improved delivery of key interventions

Between implementation and the final interventional year, the SPIN nurses saw an increasing proportion of admitted CAP cases, rising from 34.1% (FY 2013/14) to 42.0% (FY 2015/16). Associated with this was a year on year improvement in compliance with guidelines based interventions (Table 2), with all interventions implemented in >80% of admissions seen by the service. In particular, the proportion of patients receiving antibiotics within 4 hours of admission (80.6%) and dual antibiotic treatment data for severe CAP (CURB-65 score 3-5) (86.3%) exceeded reported figures for these variables from the national audit [11] (68.5% and 48.0%, respectively).

**Table 2:** CAP care bundle elements during implementation and intervention

| Source | FY | Chest x-ray within 4 hours | Antibiotics given within 4 hours | CURB-65 recorded | Dual treatment for CURB-65 3-5 cases* |
| --- | --- | --- | --- | --- | --- |
| This paper | 2013/14 | 88.8 (703) | 75.4 (596) | 42.0 (333) | 75.0 (36) |
|  | 2014/15 | 89.1 (993) | 83.3 (928) | 72.3 (805) | 87.6 (268) |
|  | 2015/16 | 91.7 (1394) | 80.6 (1227) | 93.7 (1425) | 86.3 (396) |
| Daniel et al. | 2014 | 80.2 | 68.5 | N/A | 48.0 |

\*Dual treatment calculated for CURB65 3-5 patients with drug information available (recording rate 89%).

### Improved mortality

The overall crude in-hospital and 30-day mortality rates were 16.6% and 20.1%, respectively. This was aligned with hospital episode statistics (HES) derived crude mortality rates reported by all other NHS trusts at baseline (Figure 2). Externally reported adjusted mortality rates, SHMI and HSMR, improved year on year following the implementation period. Comparing the baseline period with the intervention period (Table 3), a significant reduction in mortality was observed for both in-hospital mortality (OR=0.66 [0.60-0.73], p<0.0001) and 30-day mortality (OR=0.77 [0.70-0.85], p <0.0001) that was greatest in the subgroup of admissions seen by the SPIN nurses (in-hospital mortality: OR=0.49 [0.42-0.56], p<0.0001; 30-day mortality: OR= 0.55 [0.48-0.63], p<0.0001). A smaller but significant reduction in mortality was also observed in the intervention period for patients not seen by the SPIN service (OR=0.77 [0.69-0.86], p<0.0001). The Cox PH model demonstrated being seen by the SPIN service to have had the largest independent effect on mortality reduction (HR=0.60 [0.53-0.67], p<0.0001) (Figure 3).

**Table 3:**
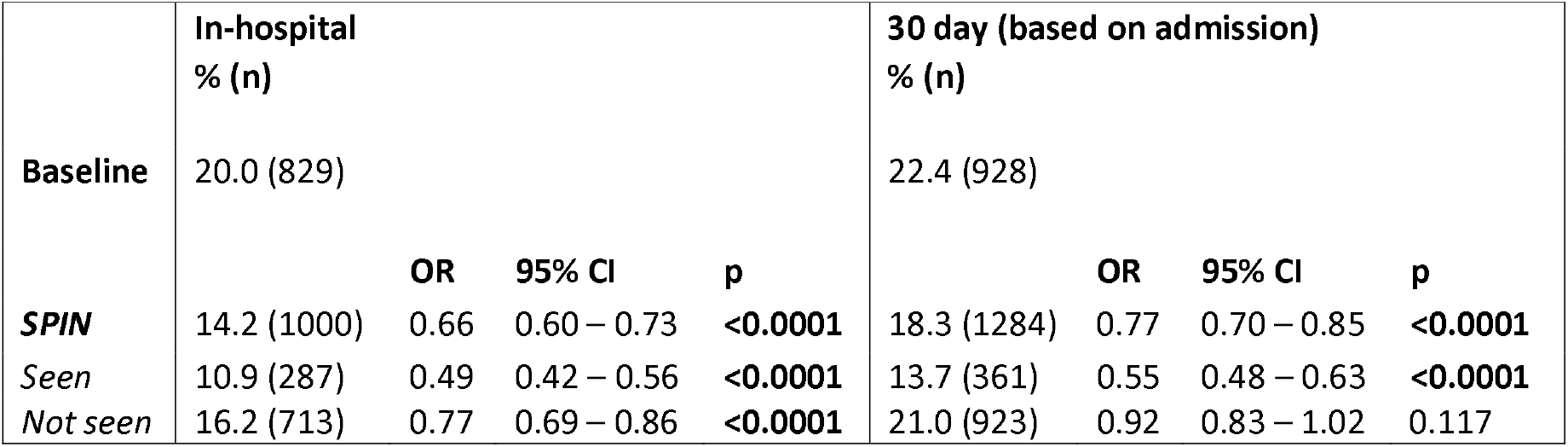
Outcomes for baseline compared to SPIN intervention period and whether patient was seen / not seen by service

|  | In-hospital<br>% (n) |  |  |  | 30 day (based on admission)<br>% (n) |  |  |  |
| --- | --- | --- | --- | --- | --- | --- | --- | --- |
| **Baseline** | 20.0 (829) |  |  |  | 22.4 (928) |  |  |  |
|  |  | **OR** | **95% CI** | **p** |  | **OR** | **95% CI** | **p** |
| **SPIN** | 14.2 (1000) | 0.66 | 0.60 – 0.73 | **<0.0001** | 18.3 (1284) | 0.77 | 0.70 – 0.85 | **<0.0001** |
| *Seen* | 10.9 (287) | 0.49 | 0.42 – 0.56 | **<0.0001** | 13.7 (361) | 0.55 | 0.48 – 0.63 | **<0.0001** |
| *Not seen* | 16.2 (713) | 0.77 | 0.69 – 0.86 | **<0.0001** | 21.0 (923) | 0.92 | 0.83 – 1.02 | 0.117 |

**Figure 2:**
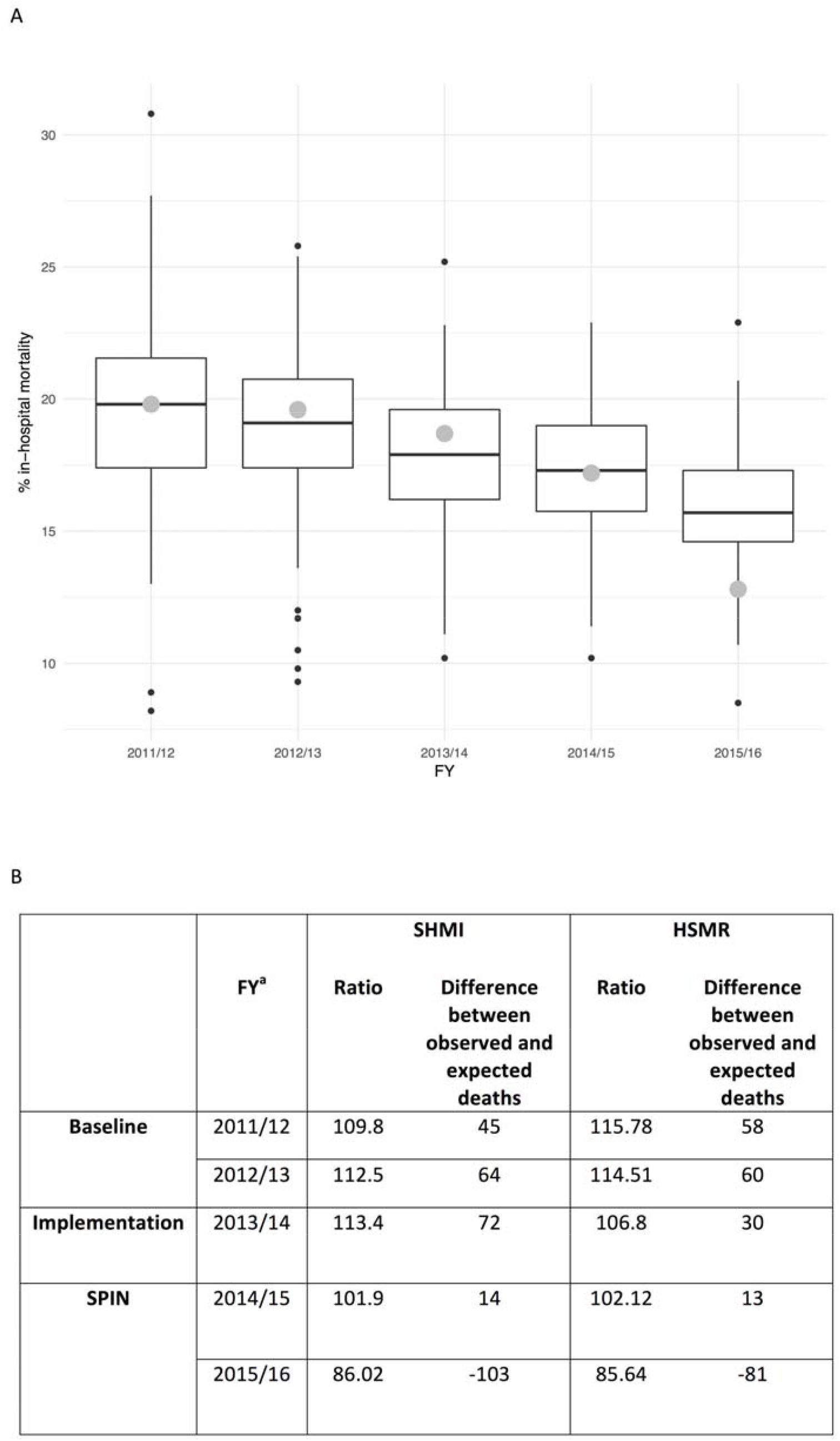
Adjusted and crude reported pneumonia related mortality. A: Crude in-hospital mortality from pneumonia admissions across all trusts B: SHMI and HSMR for all UHL pneumonia admissions. UHL mortality rates are highlighted by large grey dots. Black dots indicate outliers that are outside the interval [Q_1_ – 1.5 x IQR, Q_3_ + 1.5 x IQR] where Q_1_ = 1st quartile, Q_3_ = 3rd quartile and IQR is interquartile range Ratios are expected/observed deaths from all pneumonias over the year.

**Figure 3:**
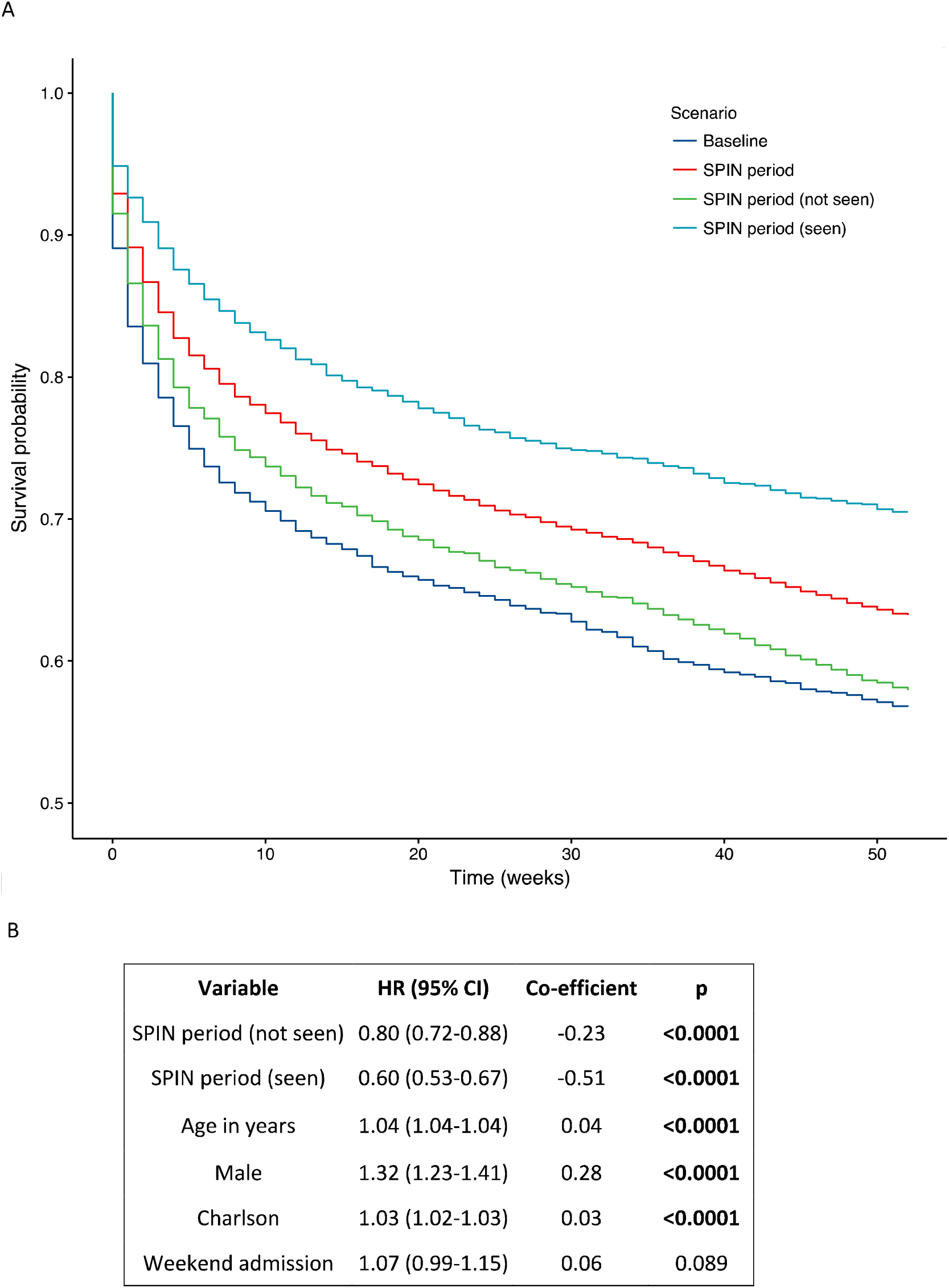
One year survival analysis comparison for CAP admissions in baseline (FY 2011/12) and SPIN (FY 2015/16) periods. A: 1-year KM survival curves for baseline and SPIN periods B: Cox PH model for 1-year survival

The median LOS for CAP admissions (6 days) did not change significantly throughout the five year period and was unaffected by the SPIN service.

## Discussion

CAP remains the most common cause of in-hospital death and the UK ranks only 21st out of 99 countries for age adjusted mortality [12]. Although better adherence to the BTS pneumonia guidelines is recognised to be associated with improved survival trends in national audits [5,11], our experience suggests there is widely inconsistent implementation of this across different departments at large NHS organisations. Reasons for this are multifactorial and include widely fluctuating seasonal admission levels for respiratory tract infections, fluctuating medical staffing levels in and out of hours and inadequate provision of resource to support services for this common but neglected condition. Here we demonstrate how, for a large and geographically disparate NHS organisation, a novel SPIN service model, comprised of only two specialist nurses working normal hours five days a week could effectively overcome this variability in the provision of care. This model achieved sustained improvements in care bundle delivery that correlated with significant reductions in crude and adjusted mortality rates for hospital admissions with CAP. In particular, our observation that the SPIN intervention appeared to reduce 30 day mortality to 13.7%, compared with the reported UK average of 17.3% [11] supports the view that suboptimal care for CAP admissions is an important driver of national mortality rates and justifies prioritisation of CAP in the recently published NHS Long Term Plan [2].

Implementation of the SPIN service improved outcome in CAP cases that were not seen by the service, indicating value and impact of education and awareness raising of CAP at a trust wide level. Given the high turnover in medical and nursing staff, sustaining this secondary benefit will require ongoing reinforcement of key messages by the service and should be viewed as an empirical component of the package of services that are provided.

There are some potential limitations to our analysis as we used a retrospective cohort design. However, it would be neither ethical nor feasible to perform a randomised prospective study to demonstrate effectiveness of these interventions for a condition known to have a poor outcome. Importantly our data confirm that rapid and robustly systematic implementation of BTS mandated standards of care significantly improve CAP prognosis, independent of disease severity, age and existing co-morbidities. The magnitude of the effect size we observed is at least equivalent to the development of new therapies for CAP and justifies the importance of improving care pathways for delivering existing resources.

Shortcomings of coded hospital episode data have been previously described, leading to concerns that CAP cases may be under represented in HES data [13,14]. However, our SPIN service model required the nurses to confirm presence of consolidation on admission chest x-rays. We believe this provides a key check point to increase the overall accuracy of pneumonia coding. To address the possibility that coding bias was introduced by adoption of the SPIN service, we compared our local hospital pneumonia coding frequencies with peer trusts and rates recorded across the UK, using standardised stationary series and found our data to be concordant.

Selection bias may have been introduced when comparing outcomes for those seen or not seen by the SPIN team during the intervention period. Indeed, higher levels of comorbidities and increased mean age were evident in patients not reviewed by the SPIN team during the intervention period that reflects a scarcity of resource within the existing SPIN service. However, a Cox PH model that included age and CCI as independent factors demonstrated the SPIN intervention to have the strongest independent effect on mortality (Figure 3). On this basis, we intend to expand the service with recruitment of more specialist nurses, providing extended cover to facilitate more equitable access to the service for admitted CAP patients.

The national pneumonia audit focussed particularly on prescribing of dual antibiotics for cases of severe pneumonia (CURB-65 3-5). Our analysis examined this point in great detail and found significantly higher compliance with dual antibiotic prescribing compared to the national audit in patients seen by the SPIN team. UHL prescribing policy favours doxycycline over macrolides as the second antibiotic in severe pneumonia. This guidance remained consistent throughout the five year observation period.

Affecting more than 250,000 people in the UK annually, CAP continues to impact drastically on UK healthcare systems, and an ageing population is projected to lead to large increases in pneumonia admissions over the next two decades [13]. In this context we conclude relatively simple specialist nurse led interventions appear to provide an effective strategy with high impact at low cost. Wider adoption of the model across the acute care sector could help transform the outcome for large numbers of emergency admissions with this life threatening condition.

## Data Availability

Since this paper was the result of a service improvement exercise, we do not have consent to share the data used to produce the conclusions in this paper.

## Acknowledgements

We would like to thank members of the SPIN team past and present for their dedication and hard-work. We would also like to thank Derek Smith at Telstra/Dr Foster for providing hospital spell data, SHMI and HSMR values.

## Footnotes

### Contributors

RCF implemented data collection tools, carried out the data extraction and analysis and wrote the manuscript, MR provided statistical advice, data analysis and manuscript review, CP collected data, JS led the SPIN team and collected data, KH contributed to SPIN and collected data, RB contributed to data analysis and trust wide implementation, PH wrote the manuscript and provided critical review, GW conceived the SPIN approach, wrote the manuscript and is the guarantor.

### Ethical approval

The conduct and reporting of this service was discussed with the local research governance committee and in line with the Policy Framework for Health and Social Care 2017, a formal ethical review was not required.

### Transparency declaration

The manuscript’s guarantor (GW) affirms that the manuscript is an honest, accurate, and transparent account of the research being reported and that no important aspects of the research have been omitted.

The corresponding author attests that all listed authors meet authorship criteria and that no others meeting the criteria have been omitted.

